# The Population Attributable Fraction (PAF) of cases due to gatherings and groups with relevance to COVID-19 mitigation strategies

**DOI:** 10.1101/2020.03.20.20039537

**Authors:** Ellen Brooks-Pollock, Jonathan M Read, Thomas House, Graham F Medley, Matt J Keeling, Leon Danon

## Abstract

**Background:** Many countries have banned groups and gatherings as part of their response to the pandemic caused by the coronavirus, SARS-CoV-2. Although there are outbreak reports involving mass gatherings, the contribution to overall transmission is unknown.

**Methods:** We used data from a survey of social contact behaviour that specifically asked about contact with groups to estimate the Population Attributable Fraction (PAF) due to groups as the relative change in the Basic Reproduction Number when groups are prevented.

**Findings:** Groups of 50+ individuals accounted for 0.5% of reported contact events, and we estimate that the PAF due to groups of 50+ people is 5.4% (95%CI 1.4%, 11.5%). The PAF due to groups of 20+ people is 18.9% (12.7%, 25.7%) and the PAF due to groups of 10+ is 25.2% (19.4%, 31.4%)

**Interpretation:** Large groups of individuals have a relatively small epidemiological impact; small and medium sized groups between 10 and 50 people have a larger impact on an epidemic.

## INTRODUCTION

Preventing social contacts has been used worldwide in 2020 to reduce transmission of the novel coronavirus, SARS-CoV-2. Early restrictions introduced sought to limit the number of people that could meet at one time. In the UK, Scotland banned all large gatherings over 500 people in March 2020 and gatherings were to be banned in England but then the national stay-at-home order was introduced on 23 March 2020. As the first lockdown was eased, gatherings or more than 30 were banned, which was then reduced to “the rule of six” in September 2020 to prevent groups of more than six individuals meeting simultaneously.

Given knowledge of transmission mechanisms, bringing together groups of people into the same space should prove conducive for the spread of close-contact infectious diseases. Indeed, gatherings have been associated with outbreaks of communicable diseases such as measles[1], influenza[2] and meningitis[3]. Public health agencies, including the World Health Organization (WHO), have specific guidance for preventing disease outbreaks at mass gatherings[4]. Factors such as age of participant[1], zoonotic transmission and presence of animals[5], crowding[6,7], lack of sanitation[7], location and event duration[6] are associated with the reporting of mass gathering-related outbreaks.

Despite the evidence of the importance of gatherings for disease transmission from intuition and individual outbreaks, the population-level impact of different mass gathering policies has not been established. While systematic reviews have identified outbreak reports involving mass gatherings[5,6], the overall impact of mass gatherings could not be quantitatively assessed. A detailed modelling study of disease transmission in the state of Georgia, USA, found that in extreme scenarios when 25% of the population participated in a 2-day long gathering shortly before the epidemic peak, peak prevalence could increase by up to 10%. More realistic scenarios resulted in minimal population-level changes[8].

The Population Attributable Fraction (PAF) is a measure of the importance of a risk factor to disease burden or death in a population, borrowed from non-communicable disease epidemiology[9]. The PAF of a risk factor is the percentage of disease burden or mortality that can be attributed to the presence of that increased risk; an example is the PAF of lung cancer cases that are due to smoking. In previous work, we demonstrated that for infectious diseases, the PAF can be estimated as the percentage change in the Basic Reproduction Number (average number of secondary cases per infectious case in an otherwise susceptible population[10]) in the counterfactual situation where the risk factor is removed from the population[11]. Here, we use representative data on individuals’ daily social contacts, including group contacts, to estimate the Population Attributable Fraction (PAF) due to groups and gatherings.

## METHODS

### Social contact data

The Social Contact Survey (SCS) [12,13] collected data on social contacts from 5,388 participants between 2009 and 2010 in the UK. Participants were asked to enumerate other people with whom they had had contact over the course of a single day. Contacts were defined as those with whom participants had a face-to-face conversation within 3 metres and/or physically touched skin-on-skin. Participants were able report individual contacts and up to five groups of contacts, for instance church groups, weddings, large work functions or multiple contacts at work. The ‘groups’ question was designed to aid participants in reporting multiple similar contacts. Group contacts were defined in the same way as individual contacts, i.e. if a person attended a concert with 1,000 people, but only spoke to 5 people, the number of recorded group contacts would be 5. Participants were asked whether members of the group knew each other.

As well as the number of contacts, participants were asked to estimate the length of time spent with each contact or group of contacts as either: less than 10 minutes, 11-30 minutes, 31-60 minutes or over 60 minutes, the distance from home, the frequency with which the contact took place and whether it involved physical contact.

The SCS data are available to download at http://wrap.warwick.ac.uk/54273/.

### Reproduction Numbers and Population Attributable Fraction

We calculate the Basic Reproduction Number with and without groups of various sizes. For each participant *j*, we use their *j*_*k*_ contact reports to calculate their individual Reproduction Number,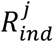. We assumed that 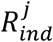. is proportional to the number of individuals reported in the contact, *n*_*i*_, (*n*_*i*_ = 1 for single contacts, *n*_*i*_ > 1 for groups) multiplied by the duration of each contact, *d*_*i*_:

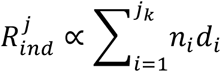

The duration of each contact is taken as the mid-point of each time interval, i.e. 5 minutes, 20 minutes, 45 minutes and 6 hours, as recorded by the participant. The interpretation of contact duration is different for individual versus group contacts, as there is a limit to the number of face-to-face contacts that one person can make in a finite time. We observe a saturation of contact duration for individuals with large numbers of contacts (figure 1B). The saturation occurs between 20 and 30 contacts per individual. We adjust for this by dividing the duration of group contacts by the number of individuals in the group, when the number of group contacts is greater than a random number between 20 and 30.

**Figure 1:**
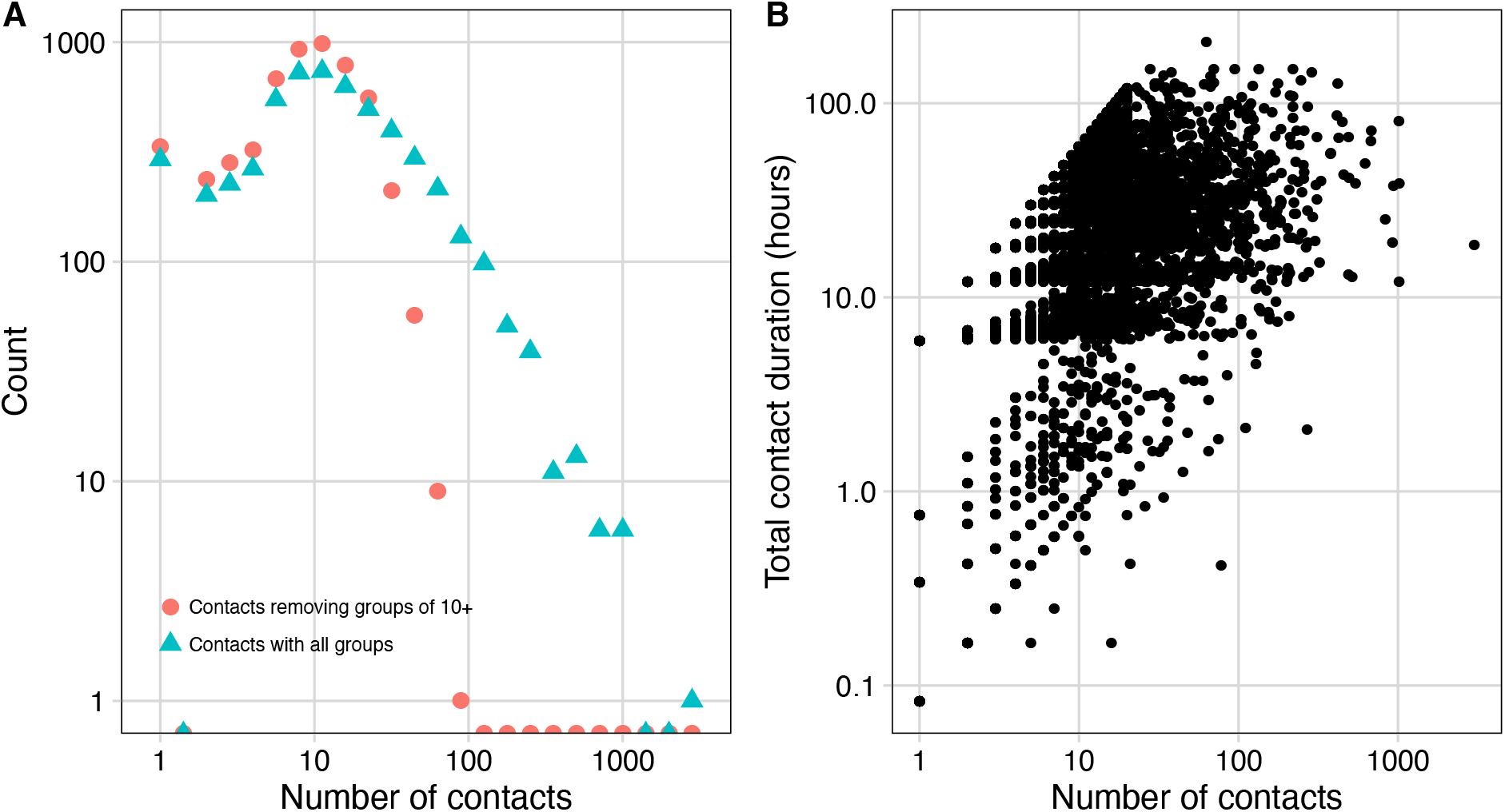
A) The distribution in the number of social contacts per participants from the Social Contact Survey (n=5,388) with and without groups of ten and greater. Even without groups of ten or more, individuals can have more than ten other contacts. B) The relationship between number of contacts and total contact duration.

The population-level reproduction number is the average number of secondary cases caused by an average infectious person. Individuals with higher 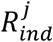 with contribute more to the population-level *R*_*t*_ because they are more likely to get infected than individuals with lower 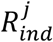. Therefore, we estimate *R*_*t*_ as a bootstrap resample (random sample with replacement) of the individual reproduction numbers weighted by the individual reproduction numbers:

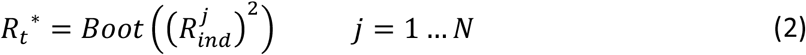

where *N* is the number of participants in the SCS. The mean and 95% Confidence Intervals were calculated from the bootstrapped sample.

We calculated the PAF for groups of size *G* or greater as the percentage change in the Basic Reproduction Number:

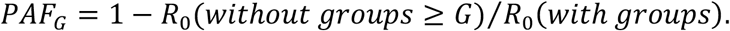

We investigate the PAF for groups of greater than 10, and up to groups greater than 100, in increments of 10. We investigated differences between groups that knew each other and groups that did not know each other.

## RESULTS

### Impact of groups on numbers of contacts per person

48,001 unique contacts were reported by 5,388 participants. Of those, 42,945 (89%) were individual contacts and 5,056 groups were reported (accounting for 11% of reported contacts). The median and mean number of contacts per person was 11.5 and 27.0, range 1 to 3,011 (figure 1A).

2,427 (45%) of participants reported group contacts. The majority of groups reported (3,860; 76%) were groups of people who knew each other. 2,979 (59%) groups had 10 or fewer members; the median and mean reported group size was 9 and 20.3 individuals respectively.

Restricting contacts to groups of size 50 or less, reduces the median and mean number of individual contacts per person to 11.0 and 18.8; restricting contacts to groups of size 20 or less, reduces the median and mean number of contacts per person to 10.0 and 14.1; restricting contacts to groups of size 10 or less, reduces the median and mean number of contacts per person to 9 and 11.0. Figure 1 shows the degree distribution (number of contacts) per person with and without contacts associated with groups of size greater than 10.

Compared to individual contacts, group contacts were more likely to be more than 2 miles from the participants home (64% versus 51%), less likely to involve physical contact (25% versus 44%) and more likely to involve new individuals (21% versus 15%).

### Population Attributable Fraction (PAF)

The PAF due to groups decreased with increasing group size. For the largest groups with more than 100 individuals the PAF100 is estimated at 0.6% (95% CI: 0.4%, 0.8%). The PAF50 is estimated at 5.5% (95% CI: 1.4%, 11.4%); the PAF20 is 18.9% (95% CI: 12.7%, 25.7%); the PAF10 is 25.2% (95% CI: 19.4%, 31.4%) (figure 2).

**Figure 2:**
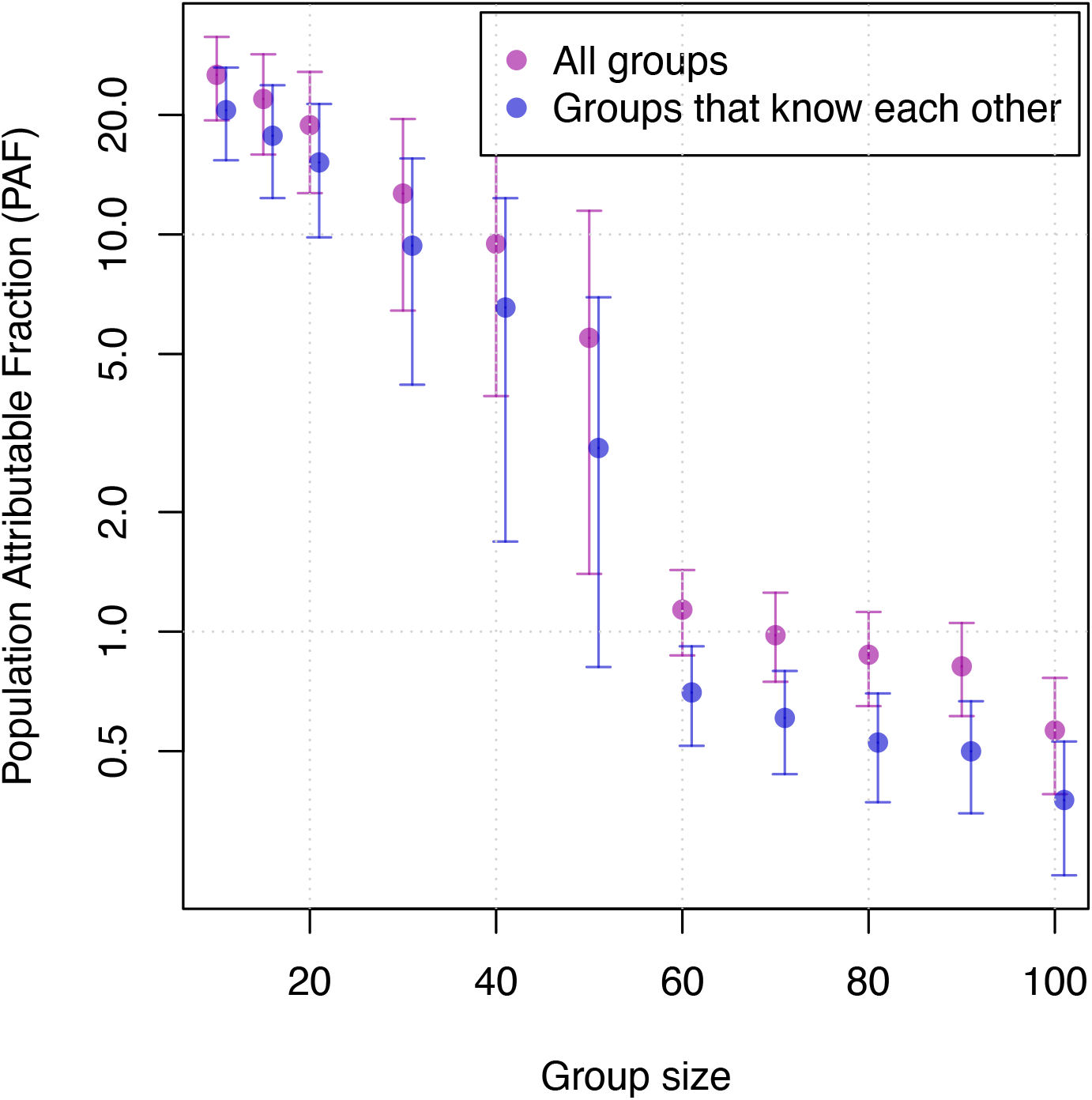
The Population Attributable Fraction (PAF) of cases due to groups of various sizes. The purple circles are all groups, the blue circles are groups of people who are known to each other. The error bars are 95% confidence intervals.

The pattern of decreasing PAF with increasing group size is seen for both groups of individuals who are known to each other and groups of individuals who are unknown to each other. The PAF due to groups of 10+ known to each other is estimated at 20.6% (95% CI: 15.4%, 26.3%) and due to groups of 50+ known to each other is estimated at 2.9% (95% CI: 0.8%, 6.9%). The remaining contribution to Rt is due to contact with individuals.

The estimated impact of large groups on *R*_*t*_ is due to the relative frequency with which they are reported in the Social Contact Survey. These results highlight the relative importance of medium-size groups of between 10 and 20 individuals.

## DISCUSSION

In this paper, we analysed social contact data in the context of infectious disease transmission and gatherings. Our findings suggest that large groups of individuals have a relatively small impact on an epidemic. This is due to the relative rarity of large-scale gatherings and the sub-linear scaling between number of contacts and infectivity.

The Social Contact Survey (SCS) is one of a number of social contact surveys that have been conducted to quantify the impact of social mixing on disease transmission[14–18]. The SCS specifically asked about groups of similar contacts. These groups are not necessarily public or mass gatherings, and represented groups that both knew each other and those that did not. The group sizes reported in the SCS were not necessarily the same size of an event where contacts may have taken place. Therefore, this analysis should be considered in terms of contacts per person, rather than to guide the acceptable size of organised events. The SCS asked about contacts on a single day and did not capture multi-day events; simulation studies have shown that prolonged mass gatherings were necessary to alter the course of an epidemic[8]. Our analysis was based on social contact data collected between 2009 and 2010; contact patterns may have altered in the past decade. We also did not account for individuals changing their behaviour if group activities were cancelled.

In the context of COVID-19 mitigation, this analysis considered one aspect of gatherings: the impact on an epidemic. However, there may be other valid reasons for preventing mass events, such as policing and managing resources. Our analysis implicitly assumes that infection is already present in the population as is the case for COVID-19; for other diseases, gatherings can be associated with increased global travel which can bring new strains into an area or result in out-of-season outbreaks, which were also not captured here.

Our findings illustrate the difficult choices that are necessary to limit COVID-19 spread. Meetings of large groups of more than 100 individuals are relatively infrequent, and their prohibition may have a limited impact on the epidemic. More epidemiologically relevant are groups of 10 to 20 people, as they occur more frequently and could potentially have a larger impact on transmission; they may also involve inter-generational family groups. This analysis was designed to aid decision-making in the context of social distancing measures to control COVID-19. and should be considered against alternative control strategies so that the most effective measures can be implemented in the long term.

## Data Availability

data are available to download at http://wrap.warwick.ac.uk/54273/.

---

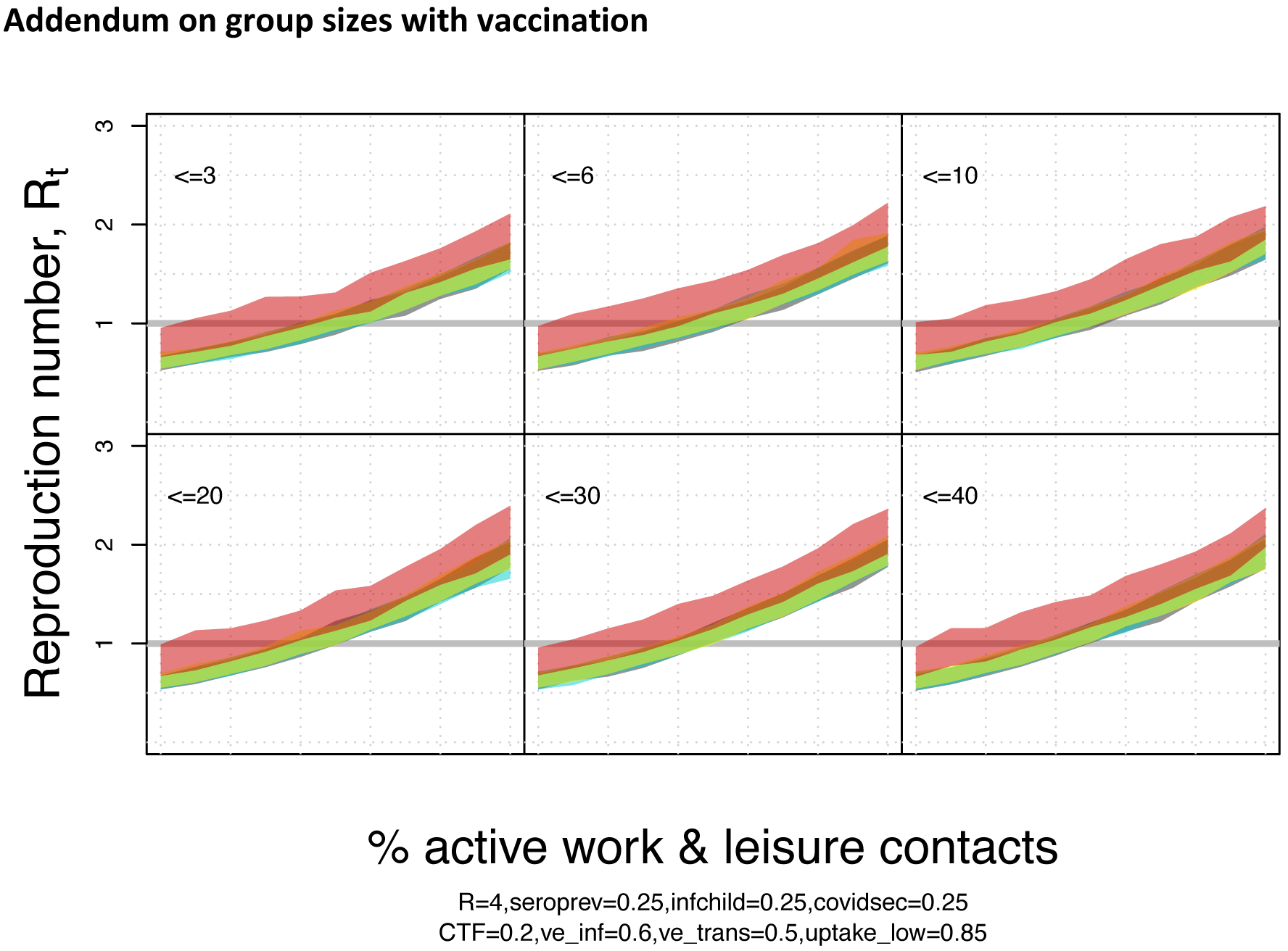

## References

[1] Hoang VT, Gautret P. Measles outbreaks at mass gathering mostly occur at youth events. Lancet Infect Dis 2020;20:23. doi:10.1016/S1473-3099(19)30685-1.

[2] Blyth CC, Foo H, van Hal SJ, Hurt AC, Barr IG, McPhie K, et al. Influenza outbreaks during world youth day 2008 mass gathering. Emerg Infect Dis 2010;16:809–15. doi:10.3201/eid1605.091136.

[3] Yezli S, Gautret P, Assiri AM, Gessner BD, Alotaibi B. Prevention of meningococcal disease at mass gatherings: Lessons from the Hajj and Umrah. Vaccine 2018;36:4603–9. doi:10.1016/j.vaccine.2018.06.030.

[4] World Health Organization (WHO). Global mass gatherings?: implications and opportunities for global health security Report by the Secretariat. vol. 86. 2011.

[5] Rainey JJ, Phelps T, Shi J. Mass gatherings and respiratory disease outbreaks in the united states-should we be worried? results from a systematic literature review and analysis of the national outbreak reporting system. PLoS One 2016;11. doi:10.1371/journal.pone.0160378.

[6] Ishola DA, Phin N. Could influenza transmission be reduced by restricting mass gatherings? Towards an evidence-based policy framework. J Epidemiol Glob Health 2011;1:33–60. doi:10.1016/j.jegh.2011.06.004.

[7] Abubakar I, Gautret P, Brunette GW, Blumberg L, Johnson D, Poumerol G, et al. Global perspectives for prevention of infectious diseases associated with mass gatherings. Lancet Infect Dis 2012;12:66–74. doi:10.1016/S1473-3099(11)70246-8.

[8] Shi P, Keskinocak P, Swann JL, Lee BY. The impact of mass gatherings and holiday traveling on the course of an influenza pandemic: A computational model. BMC Public Health 2010;10:778. doi:10.1186/1471-2458-10-778.

[9] Rosen L. An Intuitive Approach to Understanding the Attributable Fraction of Disease Due to a Risk Factor?: The Case of Smoking 2013:2932–43. doi:10.3390/ijerph10072932.

[10] Anderson R, May R. Infectious diseases of humans: dynamics and control. Oxford University Press; 1992.

[11] Brooks-Pollock E, Danon L. Defining the population attributable fraction for infectious diseases. Int J Epidemiol 2017;46:976–82. doi:10.1093/ije/dyx055.

[12] Danon L, House TA, Read JM, Keeling MJ. Social encounter networks: collective properties and disease transmission. J R Soc Interface 2012;9.

[13] Danon L, Read JM, House TA, Vernon MC, Keeling MJ. Social encounter networks: Characterizing great Britain. Proc R Soc B Biol Sci 2013;280. doi:10.1098/rspb.2013.1037.

[14] Mossong J, Hens N, Jit M, Beutels P, Auranen K, Mikolajczyk R, et al. Social contacts and mixing patterns relevant to the spread of infectious diseases. PLoS Med 2008;5:e74. doi:10.1371/journal.pmed.0050074.

[15] Hens N, Goeyvaerts N, Aerts M, Shkedy Z, Van Damme P, Beutels P. Mining social mixing patterns for infectious disease models based on a two-day population survey in Belgium. BMC Infect Dis 2009;9:5.

[16] Eames KTD, Tilston NL, Brooks-Pollock E, Edmunds WJ. Measured dynamic social contact patterns explain the spread of H1N1v influenza. PLoS Comput Biol 2012;8. doi:10.1371/journal.pcbi.1002425.

[17] Read JM, Lessler J, Riley S, Wang S, Tan LJ, Kwok KO, et al. Social mixing patterns in rural and urban areas of Southern China. Proc R Soc B Biol Sci 2014;281. doi:10.1098/rspb.2014.0268.

[18] Kwok KO, Cowling B, Wei V, Riley S, Read JM. Temporal variation of human encounters and the number of locations in which they occur: A longitudinal study of Hong Kong residents. J R Soc Interface 2018;15. doi:10.1098/rsif.2017.0838.

[19] Lemieux JE, Siddle KJ, Shaw BM, Loreth C, Schaffner SF, Gladden-Young A, et al. Phylogenetic analysis of SARS-CoV-2 in Boston highlights the impact of superspreading events. Science (80-) 2021;371. doi:10.1126/science.abe3261.

